# Personal Care Products and Incident Hypertension: Prospective Cohort Study of U.S. Women

**DOI:** 10.64898/2026.05.18.26353536

**Authors:** Jungeun Lim, Che-Jung Chang, Alexandra J. White, Gabriel A. Goodney, Hantao Wang, Jungnam Joo, Véronique L. Roger, Dale P. Sandler, Jason Y.Y. Wong

## Abstract

**Background:** Over half of U.S. women have hypertension, a strong but modifiable risk factor for cardiovascular diseases. Personal care products (PCPs) are widely used in daily life and contain endocrine disrupting chemicals that can alter hormonal regulation of blood pressure. However, the relationship between PCPs and hypertension has not been well studied. We investigated whether patterns of PCP use were associated with incident hypertension in a large prospective cohort study of U.S. women.

**Methods:** Sister Study participants were recruited in 2003–2009 and followed until September 30, 2021. Usage frequency of 41 PCPs in the 12 months before baseline was self-reported. Latent class analyses identified groups with similar PCP use patterns (“infrequent,” “moderate,” or “frequent”). At baseline, we excluded women with prevalent hypertension, antihypertensive medication users, or those missing hypertension status. Multivariable Cox regression was used to estimate associations between PCP use and incident self-reported hypertension.

**Results:** During a mean follow-up of 11.4 years, 10,099 women developed hypertension. Frequent PCP use was associated with higher hypertension risk [HR=1.08 (95% CI: 1.03, 1.13); p-trend=0.003], with a 4.1% population attributable risk. Frequent users of beauty products had higher risk than infrequent users [HR=1.11 (95% CI: 1.05, 1.16)]. Moderate and frequent users of hygiene products also had increased risk [HR=1.07 (95% CI: 1.01, 1.13); HR=1.13 (95% CI: 1.08, 1.19)].

**Conclusions:** Frequent PCP use, especially beauty and hygiene products, was associated with incident hypertension. Our findings implicate everyday chemicals as modifiable cardiovascular risk factors and highlight the need to identify pathogenic components in widely used consumer products.

## Introduction

Consumer personal care products (PCPs) contain complex mixtures of chemicals and are widely used long-term, resulting in chronic exposure to endocrine disrupting chemicals (EDCs) and other hazards. The United States (U.S.) women use more PCPs than men, applying an estimated 12 products containing 168 chemicals daily ^1^. Chronic exposure to multiple chemicals can significantly increase the risk of chronic diseases, even if individual chemicals are at low concentrations. This occurs through synergistic interactions where the combined effect of chemicals exceeds the sum of their individual effects ^2,3^. EDCs can alter circulating sex hormones ^4^, which can in turn impact the cardiovascular system, including blood pressure regulation ^5,6^.

Hypertension, a leading cardiovascular disease (CVD) risk factor, affects half of U.S. adults, with prevalence varying by age, gender, race, and socioeconomic status ^7–10^. Population studies have shown that hypertension is hormonally-responsive ^11^, with sex hormones regulating vascular function ^12^ and contributing to gender differences in blood pressure ^13^. While childhood blood pressure is similar by gender, boys have higher blood pressure compared to age-matched girls, which coincides with increased sex hormone secretion during this crucial developmental stage ^14^. Later in the life course, the influence of estrogen on blood pressure among adult women has been observed during the menstrual cycle, when blood pressure is inversely related to circulating estrogen levels ^14^.

A recent meta-analysis of epidemiologic studies showed that exposure to several types of EDCs such as per- and polyfluoroalkyl substances (PFASs) and propylparaben found in PCPs were associated with an increased risk of hypertension ^5,15^. However, the relationship between patterns of PCP use and hypertension risk has not been thoroughly explored.

We investigated the associations between patterns of PCP use and risk of incident hypertension in the Sister Study, a large prospective cohort of women residing across the U.S. We examined individual and grouped PCPs to better understand the risks associated with using multiple products in a real-world setting. Further, given potential differences in PCP use among subgroups in the general population, we also examined whether these associations varied by race and ethnicity, menopausal status, annual household income, and obesity. Identifying these everyday exposures as modifiable risk factors contributes to a better etiologic understanding of hypertension, which can help efforts to reduce the burden of cardiovascular disease.

## Methods

### Study design and population

The Sister Study is a prospective cohort study of 50,884 women aged 35 to 74 years at enrollment (2003-2009) residing in the United States, including Puerto Rico. Study design, eligibility criteria, and data collection details were reported previously ^16^. Briefly, participants completed baseline questionnaires and home visits with a trained examiner, which included measuring blood pressure, height, weight, and collecting fasting blood samples. After obtaining consent and review of self-completed forms, participants rested for several minutes. Three consecutive blood pressure measurements were taken via aneroid sphygmomanometer (model 760/ 775X; American Diagnostic Corporation). The average of the second and third systolic blood pressure (SBP) and diastolic blood pressure (DBP) measurements was used. When only two or one blood pressure measurements were available, the second measurement or the single value was used (<2%).

From the 50,884 women, we excluded those who self-reported having been diagnosed with hypertension before baseline or taking anti-hypertensive medication at enrollment (n=17,546). We also excluded women whose hypertension status was reported by next of kin or defined by National Death Index (NDI) or death certificate only (n=101). Women missing hypertension status at baseline (n=9) or the PCP questionnaire (n=2,150) were also excluded. After applying these criteria, the final analytic sample included 31,078 women. This analysis used Data Release 11.1, with follow-up through September 30, 2021. All participants provided written informed consent.

### Personal care product use

Participants self-reported their usage frequency of 41 PCPs in the 12-month period before baseline using five options: 1) did not use, 2) used less than once a month, 3) used 1-3 times per month, 4) used 1-5 times per week, and 5) used more than 5 times per week. For product group-specific analyses, these PCPs were aggregated into: twelve beauty products (blush or rouge, eye liner, shadow, mascara, foundation, lipstick, perfume or cologne, makeup remover, artificial nails/fill-ins, cuticle cream, nail polish, and nail polish remover); seven everyday hair products (hair conditioner/rinse, hair food, hair spray, hair styling gel/mousse, Minoxidil or Rogaine, pomade or hair grease, and shampoo); eight hygiene products [bath/shower gel, deodorant/antiperspirant, douche, mouthwash/rinse, shaving creams/gels, and Talcum powder (under arms, on vaginal area, or to other areas)]; and fourteen skincare products (anti-aging/wrinkle products, age spot lightener, baby/mineral-based oils, blemish/acne products, body lotions or creams, cleansing cream, face creams/moisturizers, facial masks, foot creams or moisturizers, hand lotions or creams, lip moisturizers, petroleum jelly, skin lighteners, and self-tanning products) ^17^.

### Statistical analysis

Participants who self-reported a diagnosis of hypertension during the follow-up period were defined as incident cases. We conducted two analyses to examine the association between PCP use and hypertension risk. First, we analyzed overall PCPs combined. Second, we analyzed each PCP group separately.

In the overall PCP analyses, missing values in the PCP variables and baseline characteristics were imputed using all PCP variables, baseline characteristics (i.e., SBP, DBP, family history of hypertension, age at entry, educational attainment, alcohol use, race and ethnicity, annual household income, smoking status, smoking pack-year menopausal status, electronic cigarette use, body mass index (BMI; kg/m^2^), and physical activity), and indicator for hypertension and the follow-up time estimated using the Nelson-Aalen estimator ^18^. We used the “MICE” package in R (version 4.4.1, R Foundation for Statistical Computing Vienna, Austria) and 10 imputed data sets were considered. Please see Table 2 for detailed information on imputation.

After imputation, we used random survival forest (RSF) to identify key PCPs that predict the risk of hypertension considering potential non-linear and higher order interactions between PCP variables. To determine the number of variables for the latent class analysis (LCA), a nested RSF based on the variable importance ranking was considered. In a nested RSF, the improvement in C-statistics plateaued after 33 variables (eFigure 1 in Supplement). Among the 41 PCPs, a total of 33 PCPs including 11 beauty, 6 everyday hair, 5 hygiene, and 11 skincare products were selected (eTable 1 in Supplement). eTable 2 showed the model fit indices for LCA of overall product use. We considered not only the significant decrease in AIC and BIC values, but also the substantive theory and interpretability to enhance the validity and interpretability of the model. A detailed definition of the latent classes of overall PCP usage was described in eTable 3.

**Figure 1.**
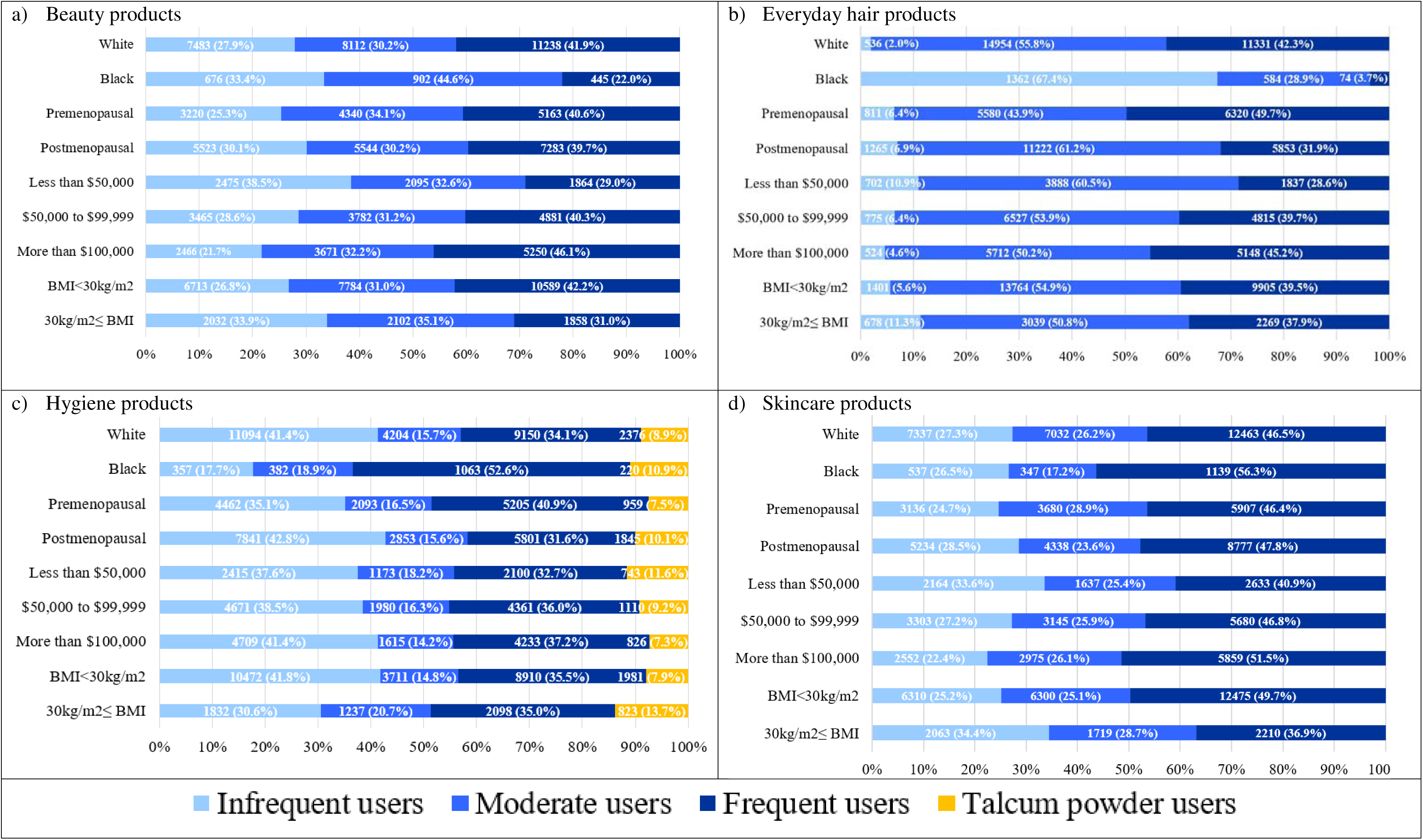
Frequency of personal care product use by race, menopausal status, annual household income, and obesity status a) Beauty products, b) Everyday hair products, c) Hygiene products, and d) Skincare products. Subjects with missing data for all products in each group (Twenty two subjects for everyday hair products, 14 subjects for hygiene products, and 1 subject for skincare products) were excluded.

For the PCP variables selected from RSF, except the no use category, we grouped adjacent categories with low frequencies (<5% of all participants). Subsequently, we performed LCA using PROC LCA in SAS statistical software (version 9.4; SAS Institute Inc.) to identify groups of individuals with similar PCP use patterns across PCP groups ^19^. The detailed methods used to define latent classes have been described ^20^. In the LCA, the best fit model was selected by comparing the Akaike’s information criterion (AIC), the Bayesian information criterion (BIC), G^2^ statistics, and theoretical interpretability ^20^.

The associations between latent classes of PCP use and the risk of hypertension were assessed using multivariable Cox proportional hazards regression models, controlling for age (years) as a time scale, and adjusting for BMI, race and ethnicity, education level, annual household income, menopausal status at enrollment, smoking pack-year, alcohol consumption, and physical activity. Pack-years of smoking were calculated by multiplying the number of packs of cigarettes smoked per day by the number of years smoked. In addition, we conducted additional stratified analyses by race and ethnicity, menopausal status, annual household income, and obesity status at baseline. Multiplicative interactions were assessed through likelihood ratio tests by comparisons of Cox models with and without the cross-product of each factor and latent class. P-values for trend across ordered latent classes were calculated. Furthermore, we examined the associations between individual PCPs and incident hypertension using multivariable Cox proportional hazards regression models with the same covariate adjustment described above. Population attributable risk (PAR) of hypertension associated with overall PCP use was estimated using Chen and Cox’s Methods ^21^. The proportional hazards assumption was assessed using Schoenfeld residuals plots and tests.

## Results

### Study population characteristics

We analyzed 31,078 women, among whom, 10,099 developed hypertension during a mean follow-up of 11.4 years (Table 1). Among participants, 6.5% self-reported their race and ethnicity as non-Hispanic Black, 86.3% as non-Hispanic White, and less than 8% as Hispanic or other. Fifty-nine percent of the study participants were postmenopausal women.

**Table 1.** Characteristics of the Sister Study participants.

| Characteristic | Overall (n=31,078) |
| --- | --- |
| Age at entry, mean (SD), y | 53.77 (8.72) |
| Race and ethnicity, n (%) |  |
| Non-Hispanic White | 26,833 (86.34) |
| Non-Hispanic Black | 2,023 (6.51) |
| Hispanic | 1,429 (4.60) |
| Other | 786 (2.53) |
| Missing | 7 (0.02) |
| Body mass index (kg/m<sup>2</sup>), mean (SD) | 26.14 (5.25) |
| Systolic blood pressure<sup>a</sup> | 111.61 (12.35) |
| Diastolic blood pressure<sup>b</sup> | 70.98 (8.32) |
| Smoking pack-years (py)<sup>c</sup> | 5.48 (11.09) |
| Smoking status, n (%) |  |
| Never smoker | 17,977 (57.84) |
| Former smoker | 10,615 (34.16) |
| Current smoker | 2,478 (7.97) |
| Missing | 8 (0.03) |
| Alcohol use, n (%) |  |
| Never or past | 5,037 (16.21) |
| Current <1 drink | 10,499 (33.78) |
| Current ≥1 drink | 15,504 (49.89) |
| Missing | 38 (0.12) |
| Post-menopause, n (%) | 18,350 (59.04) |
| Educational attainment, n (%) |  |
| High school or less | 4,164 (13.40) |
| Some college | 5,634 (18.13) |
| College and above | 21,273 (68.45) |
| Missing | 7 (0.02) |
| Annual household income, n (%) |  |
| Less than \$50,000 | 6,434 (20.70) |
| \$50,000 to \$99,999 | 12,128 (39.02) |
| More than \$100,000 | 11,387 (36.64) |
| Missing | 1,129 (3.63) |
| Physical activity, mean (SD), hours per week | 14.24 (7.95) |
| Family history of hypertension (yes), n (%)<sup>d</sup> | 20,010 (73.07) |

<sup>a</sup>Average systolic blood pressure after cleaning for implausible values (first reading excluded in presence of additional readings). One hundred seven subjects missing this variable
<sup>b</sup>Average diastolic blood pressure after cleaning for implausible values (first reading excluded in presence of additional readings). One hundred eight subjects missing this variable
<sup>c</sup> Among ever smokers
<sup>d</sup> Biological parents; There were 3,693 (11.88%) subjects with missing values for family history of hypertension.

**Table 2.** Association between overall personal care product use and hypertension risk^a^.

|  | All groups of products |  |  |  |
| --- | --- | --- | --- | --- |
|  | Infrequent users | Moderate users | Frequent users | P-trend |
| HR (95%CI)<sup>b</sup> | 1 (Ref.) | 1.02 (0.97-1.07) | 1.08 (1.03-1.13) | 0.003 |

HR=Hazard ratios; CI=95% confidence intervals; Ref.=reference group
<sup>a</sup> Missing values in the personal care product variables and participants' baseline characteristics were imputed using all personal care product variables, baseline characteristics (i.e., systolic blood pressure, diastolic blood pressure, family history of hypertension, age at entry, educational attainment, alcohol use, race and ethnicity, annual household income, smoking status, smoking pack-year menopausal status, electronic cigarette use, body mass index, and physical activity), and indicator for hypertension and the follow-up time estimated using the Nelson Aalen estimator. We used the "MICE" packages in R (version 4.4.1, R Foundation for Statistical Computing Vienna, Austria) and 10 imputed data sets were considered. The proportion of missing values in personal care product variables was relatively low (all <0.7% except one variable with 1.1% missing). We observed minimal between-imputation variability in the estimated hazard ratios for the association between personal care product variables and incident hypertension (on the order of $<10^{-6}$ for all personal care products). Accordingly, we summarized the results using the mode across the 10 imputations.
<sup>b</sup> Models accounted for age as the timescale. In addition, models were adjusted for race/ethnicity (non-Hispanic Black, non-Hispanic White, Hispanic, or other,), body mass index (continuous, kg/m<sup>2</sup>), education level (high school or less, some college, or college and above), annual household income (<\$50,000, \$50,000-<\$100,000, ≥\$100,000), menopausal status at enrollment (premenopausal or postmenopausal), smoking pack-year (continuous, pack-year), alcohol consumption (never or past, current <1 drink, or current ≥1 drinks), and physical activity (continuous, hours per week) after imputation of missing values (<5%) as described in the methods section.

### PCP usage patterns by race and ethnicity, menopausal status, and annual household income

For LCA, original response options in the questionnaire were grouped into three (‘infrequent’, ‘moderate’, and ‘frequent’) for beauty, everyday hair, and skincare products and into four for hygiene products (adding ‘talcum powder user’).

Compared to White women, Black women more frequently used hygiene (52.6% vs. 34.1%) and skincare products (56.3% vs. 46.5%), but less frequently used beauty (22.0% vs. 41.9%) and everyday hair products (3.7% vs. 42.3%, Figure 1). Except for skincare products, premenopausal women showed higher frequent use of beauty, everyday hair, and hygiene products than post-menopausal women (40.6%, 49.7%, 40.9% vs. 39.7%, 31.9%, 31.6%, respectively). Additionally, groups with higher income levels had a higher proportion of women who frequently used PCPs, except for hygiene products. Obese women (BMI≥30kg/m^2^) used beauty, everyday hair, and skincare products less frequently than non-obese women (BMI<30kg/m^2^), but used hygiene products more frequently.

### Overall PCP use and risk of incident hypertension

We found a positive exposure-response relationship between overall PCP use and risk of hypertension, even after accounting for confounders [frequent users, HR = 1.08 (95%CI:1.03,1.13); P-trend = 0.003] (Table 2). We found some evidence of heterogeneity in the overall PCP findings by race and ethnicity, with higher hypertension risks observed only among White women (eTable 4 in Supplement). However, interactions between race and ethnicity and PCP use were non-significant (P for interaction = 0.167; eTable 4 in Supplement). In analyses stratified by menopausal status, frequent PCP users had a higher hypertension risk compared to infrequent users in both premenopausal [HR = 1.09 (95%CI:1.001,1.19); P-trend = 0.034] and postmenopausal women [HR = 1.07 (95%CI:1.01,1.13); P-trend = 0.033] (eTable 4 in Supplement). When examining socioeconomic status, a significant positive association was observed between frequent PCP use and the risk of hypertension only among women with annual household incomes under $50,000, but there were no significant interactions between income and PCP use on hypertension (P for interaction = 0.311). Lastly, even though the overall PCP-hypertension association was observed only among women with BMI <30 kg/m^2^ (p-trend_ordinal_=0.011), we did not find evidence for interaction by obesity status (P for interaction = 0.723).

### PCP groups and risk of incident hypertension

We found a positive exposure-response relationship between use of beauty products and hypertension risk [HR_frequent versus infrequent_ =1.11 (95%CI:1.05,1.16); P trend<0.001; Table 3]. In addition, a similarly strong exposure-response relationship was observed for hygiene products [HR_moderate versus infrequent_=1.07 (95%CI:1.01,1.13); HR_frequent versus infrequent_=1.13 (95%CI:1.08,1.19); P-trend<0.0001]. However, we did not detect positive associations between everyday hair product or skincare product use and hypertension risk (Table 3). The results of the sensitivity analysis that includes additional adjustment for family history of hypertension were consistent with the results of the main analysis (eTable 5 in Supplement).

**Table 3.** Associations between personal care product groups and risk of hypertension.

|  | Infrequent users | Moderate users | Frequent users | Talcum powder users | P-trend |
| --- | --- | --- | --- | --- | --- |
| **Beauty products** |  |  |  |  |  |
| Cases/n<sup>a</sup> | 2,972/8,745 | 3,149/9,886 | 3,978/12,447 | – |  |
| HR (95% CI)<sup>b</sup> | 1 (Ref.) | 1.02 (0.97-1.07) | 1.11 (1.05-1.16) | – | <0.001 |
| **Everyday hair products** |  |  |  |  |  |
| Cases/n<sup>a</sup> | 833/2,079 | 5,578/16,803 | 3,679/12,174 | – |  |
| HR (95% CI)<sup>b</sup> | 1 (Ref.) | 0.97 (0.89-1.07) | 0.99 (0.89-1.09) | – | 0.739 |
| **Hygiene products** |  |  |  |  |  |
| Cases/n<sup>a</sup> | 3,693/12,304 | 1,707/4,948 | 3,595/11,008 | 1,100/2,804 |  |
| HR (95% CI)<sup>b</sup> | 1 (Ref.) | 1.07 (1.01-1.13) | 1.13 (1.08-1.19) | 1.18 (1.11-1.27) | <0.001 |
| **Skincare products** |  |  |  |  |  |
| Cases/n<sup>a</sup> | 2,951/8,373 | 2,614/8,019 | 4,533/14,685 | – |  |
| HR (95% CI)<sup>b</sup> | 1 (Ref.) | 1.02 (0.96-1.07) | 0.99 (0.94-1.04) | – | 0.607 |

HR=Hazard ratios; CI=95% confidence intervals; Ref.=reference group
<sup>a</sup> Numbers of total subjects and hypertension events are for women with complete data for each product class and covariates only
<sup>b</sup> Models accounted for age as the timescale. In addition, models were adjusted for race/ethnicity (non-Hispanic Black, non-Hispanic White, Hispanic, other, or missing), body mass index (continuous, kg/m<sup>2</sup>), education level (high school or less, some college, college and above, or missing), annual household income (<\$50,000, \$50,000-<\$100,000, ≥\$100,000), menopausal status at enrollment (premenopausal, postmenopausal, or missing), smoking pack-year (continuous, pack-year), alcohol consumption (never or past, current <1 drink, current ≥1 drinks, or missing), and physical activity (continuous, hours per week).

In analyses stratified by race and ethnicity, menopausal status, annual household income, and obesity status, a positive association between use of beauty products and risk of hypertension was consistently observed in all strata except in Black women and obese women with BMI≥30kg/m^2^ (Table 4). A positive association was also observed between hygiene product use and risk of hypertension in the same groups. Moderate use of skincare products was associated with increased risk of hypertension among the lowest income group [HR=1.13 (95%CI:1.02,1.26)], but the association was attenuated towards the null for the most frequent users.

**Table 4.** Personal care product groups and risk of hypertension stratified by race, menopausal status, and annual household income^a^.

|  | Infrequent users | Moderate users | Frequent users | Talcum powder users | P-trend | P for interaction<sup>b</sup> |
| --- | --- | --- | --- | --- | --- | --- |
|  | HR (95%CI) | HR (95%CI) | HR (95%CI) |  |  |  |
| **Beauty products** |  |  |  |  |  |  |
| Race |  |  |  |  |  |  |
| White | 1 (Ref.) | 1.003 (0.95-1.06) | 1.11 (1.05-1.17) | – | <0.0001 | 0.241 |
| Black | 1 (Ref.) | 1.12 (0.95-1.31) | 1.19 (0.98-1.44) | – | 0.0689 |  |
| Menopausal status |  |  |  |  |  |  |
| Premenopausal | 1 (Ref.) | 1.02 (0.94-1.12) | 1.10 (1.01-1.21) | – | 0.020 | 0.681 |
| Postmenopausal | 1 (Ref.) | 1.02 (0.95-1.08) | 1.10 (1.04-1.17) | – | 0.001 |  |
| Income |  |  |  |  |  |  |
| Less than \$50,000 | 1 (Ref.) | 1.09 (0.99-1.20) | 1.13 (1.02-1.24) | – | 0.014 | 0.235 |
| \$50,000 to \$99,999 | 1 (Ref.) | 0.97 (0.90-1.05) | 1.07 (0.995-1.16) | – | 0.048 | |
| More than \$100,000 | 1 (Ref.) | 1.02 (0.92-1.12) | 1.12 (1.02-1.23) | – | 0.009 | |
| Obesity |  |  |  |  |  |  |
| Body mass index<30kg/m<sup>2</sup> | 1 (Ref.) | 1.00 (0.94-1.06) | 1.10 (1.04-1.16) | – | 0.001 | 0.491 |
| 30kg/m<sup>2</sup>≤ Body mass index | 1 (Ref.) | 1.03 (0.94-1.13) | 1.09 (0.99-1.19) | – | 0.083 |  |
| **Everyday hair products** |  |  |  |  |  |  |
| Race and ethnicity |  |  |  |  |  |  |
| White | 1 (Ref.) | 1.07 (0.92-1.24) | 1.10 (0.94-1.27) | – | 0.135 | 0.413 |
| Black | 1 (Ref.) | 0.98 (0.84-1.15) | 0.83 (0.56-1.23) | – | 0.468 |  |
| Menopausal status |  |  |  |  |  |  |
| Premenopausal | 1 (Ref.) | 0.91 (0.76-1.09) | 0.93 (0.77-1.12) | – | 0.959 | 0.137 |
| Postmenopausal | 1 (Ref.) | 1.00 (0.89-1.12) | 1.01 (0.90-1.14) | – | 0.654 |  |
| Income |  |  |  |  |  |  |
| Less than \$50,000 | 1 (Ref.) | 0.96 (0.83-1.12) | 0.96 (0.81-1.13) | – | 0.680 | 0.583 |
| \$50,000 to \$99,999 | 1 (Ref.) | 0.91 (0.77-1.07) | 0.91 (0.77-1.08) | – | 0.660 | |
| More than \$100,000 | 1 (Ref.) | 1.08 (0.88-1.33) | 1.15 (0.93-1.43) | – | 0.058 | |
| Obesity |  |  |  |  |  |  |
| Body mass index<30kg/m<sup>2</sup> | 1 (Ref.) | 1.01 (0.90-1.13) | 1.03 (0.91-1.16) | – | 0.441 | 0.037 |
| 30kg/m<sup>2</sup>≤ Body mass index | 1 (Ref.) | 0.91 (0.77-1.08) | 0.91 (0.75-1.09) | – | 0.477 |  |
| **Hygiene products** |  |  |  |  |  |  |
| Race and ethnicity |  |  |  |  |  |  |
| White | 1 (Ref.) | 1.07 (1.00-1.13) | 1.14 (1.09-1.20) | 1.17 (1.09-1.26) | <0.001 | 0.985 |
| Black | 1 (Ref.) | 1.06 (0.84-1.34) | 1.12 (0.92-1.36) | 1.15 (0.88-1.49) | 0.212 |  |
| Menopausal status |  |  |  |  |  |  |
| Premenopausal | 1 (Ref.) | 1.05 (0.95-1.17) | 1.15 (1.06-1.25) | 1.27 (1.12-1.44) | <0.001 | 0.145 |
| Postmenopausal | 1 (Ref.) | 1.08 (1.01-1.16) | 1.13 (1.06-1.19) | 1.14 (1.05-1.24) | <0.001 |  |
| Income |  |  |  |  |  |  |
| Less than \$50,000 | 1 (Ref.) | 1.05 (0.94-1.18) | 1.14 (1.04-1.26) | 1.22 (1.07-1.38) | <0.001 | 0.922 |
| \$50,000 to \$99,999 | 1 (Ref.) | 1.06 (0.96-1.16) | 1.12 (1.04-1.20) | 1.20(1.07-1.33) | <0.001 | |
| More than \$100,000 | 1 (Ref.) | 1.09 (0.98-1.22) | 1.15 (1.06-1.25) | 1.11 (0.97-1.27) | 0.002 | |
| Obesity |  |  |  |  |  |  |
| Body mass index<30kg/m<sup>2</sup> | 1 (Ref.) | 1.06 (0.99-1.14) | 1.14 (1.08-1.20) | 1.20 (1.10-1.30) | <0.001 | 0.125 |
| 30kg/m<sup>2</sup>≤ Body mass index | 1 (Ref.) | 1.05 (0.94-1.16) | 1.09(0.99-1.19) | 1.07 (0.95-1.21) | 0.102 |  |
| **Skincare products** |  |  |  |  |  |  |
| Race and ethnicity |  |  |  |  |  |  |
| White | 1 (Ref.) | 1.00 (0.95-1.06) | 0.99 (0.94-1.04) | – | 0.531 | 0.153 |
| Black | 1 (Ref.) | 1.12 (0.90-1.39) | 1.11 (0.94-1.31) | – | 0.242 |  |
| Menopausal status |  |  |  |  |  |  |
| Premenopausal | 1 (Ref.) | 1.03 (0.92-1.10) | 1.02 (0.94-1.11) | – | 0.624 | 0.619 |
| Postmenopausal | 1 (Ref.) | 1.03 (0.96-1.10) | 0.97 (0.92-1.03) | – | 0.298 |  |
| Annual household income |  |  |  |  |  |  |
| Less than \$50,000 | 1 (Ref.) | 1.13 (1.02-1.26) | 1.07 (0.98-1.18) | – | 0.151 | 0.008 |
| \$50,000 to \$99,999 | 1 (Ref.) | 0.94 (0.86-1.02) | 0.93 (0.86-1.002) | – | 0.067 | |
| More than \$100,000 | 1 (Ref.) | 0.99 (0.90-1.09) | 0.97 (0.89-1.06) | – | 0.476 | |
| Obesity |  |  |  |  |  |  |
| Body mass index<30kg/m<sup>2</sup> | 1 (Ref.) | 1.01 (0.95-1.08) | 0.97 (0.92-1.03) | – | 0.260 | 0.237 |
| 30kg/m<sup>2</sup>≤ Body mass index | 1 (Ref.) | 1.00 (0.91-1.09) | 1.03 (0.94-1.13) | – | 0.491 |  |

HR=Hazard ratios; CI=95% confidence intervals; Ref.=reference group
<sup>a</sup>Models accounted for age as the timescale. In addition, models were adjusted for race/ethnicity (non-Hispanic Black, non-Hispanic White, Hispanic, other, or missing), body mass index (continuous, kg/m<sup>2</sup>), education level (high school or less, some college, college and above, or missing), annual household income (<\$50,000, \$50,000- <\$100,000, ≥\$100,000), menopausal status at enrollment (premenopausal, postmenopausal, or missing), smoking pack-year (continuous, pack-year), alcohol consumption (never or past, current <1 drink, current ≥1 drinks, or missing), and physical activity (continuous, hours per week)
<sup>b</sup>Interactions were assessed through likelihood ratio tests by comparisons of Cox models with and without the cross-product of each factor and latent class.

### Use of individual PCPs and risk of incident hypertension

Among the 12 individual beauty products examined, usage frequency of eye shadow, foundation makeup, perfume or cologne, artificial nails or fill-ins, lipstick, eye mascara, or makeup remover were associated with an increased risk of hypertension (all FDR adjusted P-trend<0.01; eTable 6 in Supplement). For the individual everyday hair products, frequent users of hair spray had higher risk of hypertension compared with non-users (FDR adjusted P-trend<0.001). Compared to non-users, weekly hair gel/mousse use (1–5 times) showed a higher risk of hypertension [HR=1.06 (95%CI: 1.003, 1.12)], though the trend was non-monotonic (eTable 7 in Supplement). For individual hygiene products, all eight hygiene products were positively associated with hypertension risk (FDR adjusted P-trend<0.05, eTable 8 in Supplement). The strongest association was observed between douche use and risk of hypertension, with a HR of 1.29 (95%CI:0.61,2.71), in the group using it more than 5 times per week. Among the 14 individual skincare products, foot cream or lip moisturizer use was positively associated with hypertension risk (P-trend<0.05), but the significance disappeared after FDR adjustment (eTable 9 in Supplement).

## Discussion

### Findings and Interpretation

In a large prospective cohort of U.S. women, frequent use of PCPs, especially beauty and hygiene products, was associated with an increased risk of incident hypertension, even after accounting for confounders and known risk factors. This is the first study linking incident hypertension to commonly used PCPs. Given their widespread use of PCPs in daily life, especially among women, our findings have important implications for cardiovascular health in the general U.S. population.

Supporting our findings, previous epidemiologic studies have suggested a link between exposure to individual EDCs, which are commonly found in PCP, and risk of hypertension. A meta-analysis of 13 studies showed that perfluorononanoic acid (PFNA), perfluorooctanoic acid (PFOA), perfluorooctane sulfonate (PFOS), and perfluorohexane sulfonic acid (PFHxS) were significantly associated with hypertension risk ^5^. Similarly, National Health and Nutrition Examination Surveys (NHANES) 2011-2012 data linked higher urinary concentration of phthalates and heavy metals to high blood pressure (≥140 mmHg SBP and ≥90 mmHg DBP) ^22^. Conversely, a cohort study within a sub-cohort of Granada EPIC-Spain center evidenced no associations between serum concentrations of non-persistent environmental pollutants such as four parabens and bisphenol A with arterial hypertension, with the exception of an increased risk of arterial hypertension observed in the highest propylparaben percentiles propylparaben ^15^. The authors emphasized that their use of one spot serum sample for exposure characterization may not have accurately reflected the exposure to non-persistent environmental pollutants. Our prospective analysis of comprehensive questionnaire data on the use of various types of PCPs used routinely in daily life may reflect chronic exposure to complex chemical mixtures.

We found that usage frequency of beauty and hygiene products was associated with an increased risk of hypertension, whereas everyday hair and skincare products was not. We do not have any information on specific products or ingredients in this analysis; however, common everyday hair and skincare products used in the U.S. contain EDCs such as phthalates, parabens, and phenols ^23,24^. In particular, hair products marketed to Black women, such as hot oil treatment, anti-frizz/polish, leave-in conditioner, root stimulator, hair lotion, and relaxer, contain higher level of these compounds compared to other hair products ^23,24^. The everyday hair products included in this analysis may not have fully captured hair products used by Black women. For example, styles like braids or straightening often involve less frequent shampooing. Further studies including a wider range of hair products and diverse populations are needed. Regarding skincare products, although the specific EDC types and concentrations can vary depending on the product formulation, skincare products potentially have substantial endocrine disruptors due to prolonged skin contact and deeper penetration ^25^.

### Possible Explanation of the Association

Although hypertension is a known hormonally-responsive disease ^11^ the pathobiological mechanisms of EDCs remain unclear. Animal studies indicate that exposure to abnormal levels of endogenous or environmental estrogens increases the risk of developing hypertension through alteration of ion channel inhibition/activation, cardiac Ca^2+^ handling, oxidative stress, and genome/transcriptome modifications ^26^. EDCs exert agonistic and antagonistic effects on estrogen receptor ^27^. Given that cardiovascular estrogen receptors regulate vasodilation and lipids, some EDCs may affect blood pressure through these receptors ^15,28,29^. Despite the prominence of EDCs in the composition of PCPs, we cannot discount the pathogenic role of other constituents in the complex chemical mixtures or co-exposures.

### Strengths and Limitations

Our study had many strengths. In this large prospective cohort study, we not only considered the effect of individual PCPs on hypertension, but also considered the effects of the PCP latent classes, which account for patterns of use. In particular, we used latent class models, which are known to be useful tools for identifying individuals with shared real-life profiles of chemical exposures in epidemiology ^30^. Furthermore, the prospective study design ensures temporality between the exposures, covariates, and outcome, strengthening the possibility of a causal relationship.

Our investigation had some limitations. First, we analyzed a finite number of products; however, the Sister Study is among the largest existing prospective cohorts with comprehensive PCP data. Second, self-reported hypertension may underestimate actual incidence ^31^. However, studies show high concordance; for example, the Growing Up Today Study reported 85.5% agreement (kappa=0.72, 100% sensitivity, 75.3% specificity) ^32^. The China Health and Retirement Longitudinal Study also showed 81% agreement for the diagnosis of hypertension between self-report and clinical measures ^33^. Importantly, underestimating hypertension cases likely attenuates the HRs toward the null. As such, our estimates are likely conservative and the true effect is probably stronger than reported. Third, we did not have information on duration of PCP use, product ingredients, or brands. These were not asked to limit participation burden, as women may use multiple brands. Lastly, the analyses for Black women were underpowered. Given their distinct product use, chemical exposure profiles from PCPs may vary by racial and ethnic subgroups.

## Perspectives

This large-scale U.S. prospective cohort study provides evidence connecting patterns of PCP use to incident hypertension in women. While the observed HRs were modest individually, the ubiquitous and daily use of PCPs means that even small increases in risk can translate to a meaningful population-level burden. Here, an estimated 4.1% of hypertension incidence was attributable to frequent PCP use overall. Given that modest shifts in blood pressure distribution at the population level can substantially influence cardiovascular disease burden, these findings have public health significance. Future studies should incorporate biomarker-based exposure assessment to identify the specific chemical constituents responsible for the observed associations with beauty and hygiene products (e.g., phthalates, parabens, PFAS, or other endocrine disrupting chemicals). In the future, investigating whether product reformulation or exposure reduction can reduce hypertension risk could potentially provide actionable evidence for regulatory and clinical recommendations. Further research in ethnically-varied populations is also needed, as usage patterns and chemical exposure profiles vary across racial and ethnic subgroups. Finally, integrating product use data with proteomic and epigenomic profiling may elucidate the biological pathways through which chronic, low-dose chemical mixtures influence blood pressure regulation. If confirmed, our findings would support expanding the scope of modifiable cardiovascular risk factors to include everyday PCPs and could inform product safety standards aimed at reducing the cardiovascular burden partly attributable to environmental chemicals. However, caution is recommended when interpreting these observational findings and additional replication in large population cohorts is needed.

## Supporting information

All 9 supplementary tables figures

## Data Availability

All data necessary to reproduce the current analysis are publicly available upon request as described on the Sister Study website (https://sisterstudy.niehs.nih.gov/English/data-requests.htm).

## Acknowledgements

The contributions of the NIH authors were made as part of their official duties as NIH federal employees, are in compliance with agency policy requirements, and are considered Works of the United States Government. However, the findings and conclusions presented in this paper are those of the authors and do not necessarily reflect the views of the NIH or the U.S. Department of Health and Human Services. We thank Dr. Shelton Lo for his helpful input.

## CRediT authorship contribution statement

Jungeun Lim: Conceptualization, Project administration, Formal analysis, Methodology, Writing – original draft. Che-Jung Chang: Methodology, Writing – review and editing. Alexandra J. White: Writing – review and editing. Hantao Wang: Writing – review and editing. Gabriel Goodney: Writing – review and editing. Jungnam Joo: Formal analysis, Writing – review and editing. Véronique L. Roger: Writing – review and editing. Dale P. Sandler: Writing – review and editing, Methodology, Funding acquisition. Jason Y.Y. Wong: Conceptualization, Methodology, Supervision, Funding acquisition, Writing – review and editing.

## Generative Artificial Intelligence Declaration

Generative AI tools (Claude, Anthropic; ChatGPT, OpenAI) were used solely for editing and error detection. The authors take full responsibility for all content.

## Nonstandard Abbreviations and Acronyms

AIC: Akaike information criterion BIC Bayesian information criterion
CVD: cardiovascular disease
DBP: diastolic blood pressure
EDC: endocrine disrupting chemical
FDR: false discovery rate
LCA: latent class analysis
NDI: National Death Index
NHANES: National Health and Nutrition Examination Survey PAR population attributable risk
PCP: personal care product
PFAS: per- and polyfluoroalkyl substances
PFHxS: perfluorohexane sulfonic acid
PFNA: perfluorononanoic acid
PFOA: perfluorooctanoic acid
PFOS: perfluorooctane sulfonate
RSF: random survival forest
SBP: systolic blood pressure

## NOVELTY AND RELEVANCE

### What Is New?

- This is the first prospective cohort study to link patterns of personal care product use with incident hypertension in women.
- Using latent class analysis to capture real-world patterns of product use across 41 personal care products, we found that frequent users had an estimated 8% higher risk of developing hypertension compared to infrequent users.
- Beauty products and hygiene products had the most consistent associations, with exposure-response relationships observed for both.

### What Is Relevant?

- Hypertension affects half of U.S. adults and is a leading modifiable risk factor for cardiovascular disease. Women use an estimated 12 personal care products daily, resulting in chronic exposure to complex chemical mixtures, including endocrine disrupting chemicals, that can alter blood pressure regulation through hormonal pathways.
- Prior studies have examined individual chemicals (such as PFAS or phthalates) and hypertension risk, but none have evaluated how the combined pattern of everyday product use relates to developing hypertension over time.

### Clinical/Pathophysiological Implications?

- Personal care products are modifiable, everyday source of chemical exposure. Reducing frequent use of personal care products overall could prevent an estimated 4.1% of hypertension cases in U.S. women.
- Even modest increases in blood pressure at the population level can increase the burden of cardiovascular disease, making these common consumer exposures a potentially important target for public health intervention.
- These findings support further research to identify the specific chemical ingredients in beauty and hygiene products responsible for the observed associations.

---

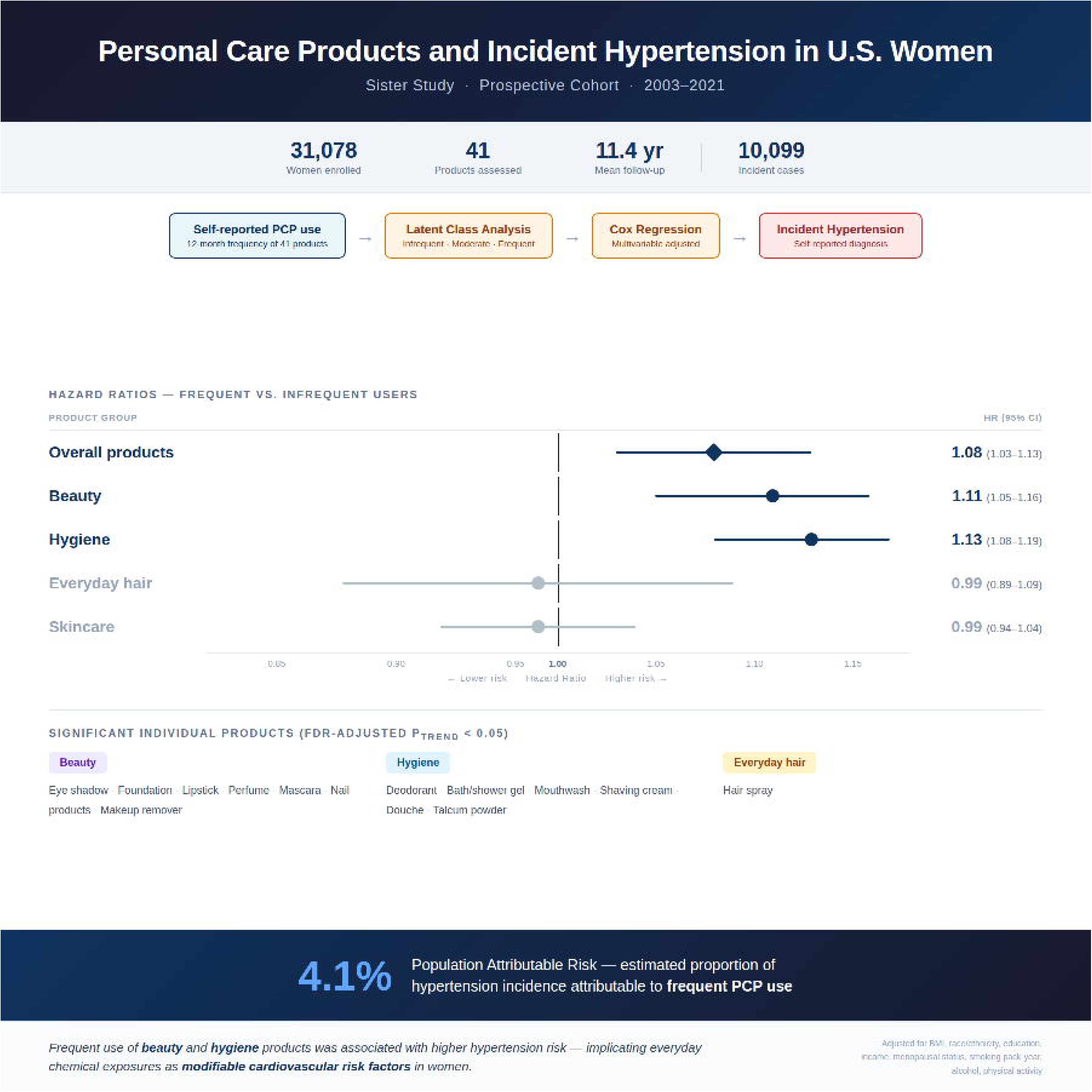

