## Supplementary material for "Personal Care Products and Incident Hypertension: Prospective Cohort Study of U.S. Women": All 9 supplementary tables figures

eTable 1. Thirty-three products selected by random survival forest.

| Ranking | Products | Product groups |
|---|---|---|
| 1 | Hair food | Hair products |
| 2 | Artificial nails (else) | Beauty products |
| 3 | Face cream | Skincare products |
| 4 | Talc (vaginal) | Hygiene products |
| 5 | Artificial nails | Beauty products |
| 6 | Douche | Hygiene products |
| 7 | Nail polish remover | Beauty products |
| 8 | Body lotion | Skincare products |
| 9 | Lip Moisturizer | Skincare products |
| 10 | Minoxidil | Hair products |
| 11 | Conditioner | Hair products |
| 12 | Lipstick | Beauty products |
| 13 | Hand lotion | Skincare products |
| 14 | Perfume | Beauty products |
| 15 | Mascara | Beauty products |
| 16 | Skin lightener | Skincare products |
| 17 | Blush | Beauty products |
| 18 | Nail polish | Beauty products |
| 19 | Talc (other) | Hygiene products |
| 20 | Deodorant | Hygiene products |
| 21 | Talc (under arms) | Hygiene products |
| 22 | Pomade | Hair products |
| 23 | Eye liner | Beauty products |
| 24 | Hair styling gel | Hair products |
| 25 | Hair spray | Hair products |
| 26 | Age spot lightener | Skincare products |
| 27 | Baby oil | Skincare products |
| 28 | Anti-aging product | Skincare products |
| 29 | Self-tanning product | Skincare products |
| 30 | Foundation | Beauty products |
| 31 | Foot cream | Skincare products |
| 32 | Cleansing cream | Skincare products |
| 33 | Bath gel | Hygiene products |

eFigure 1. Variable Importance Plot of random survival forest

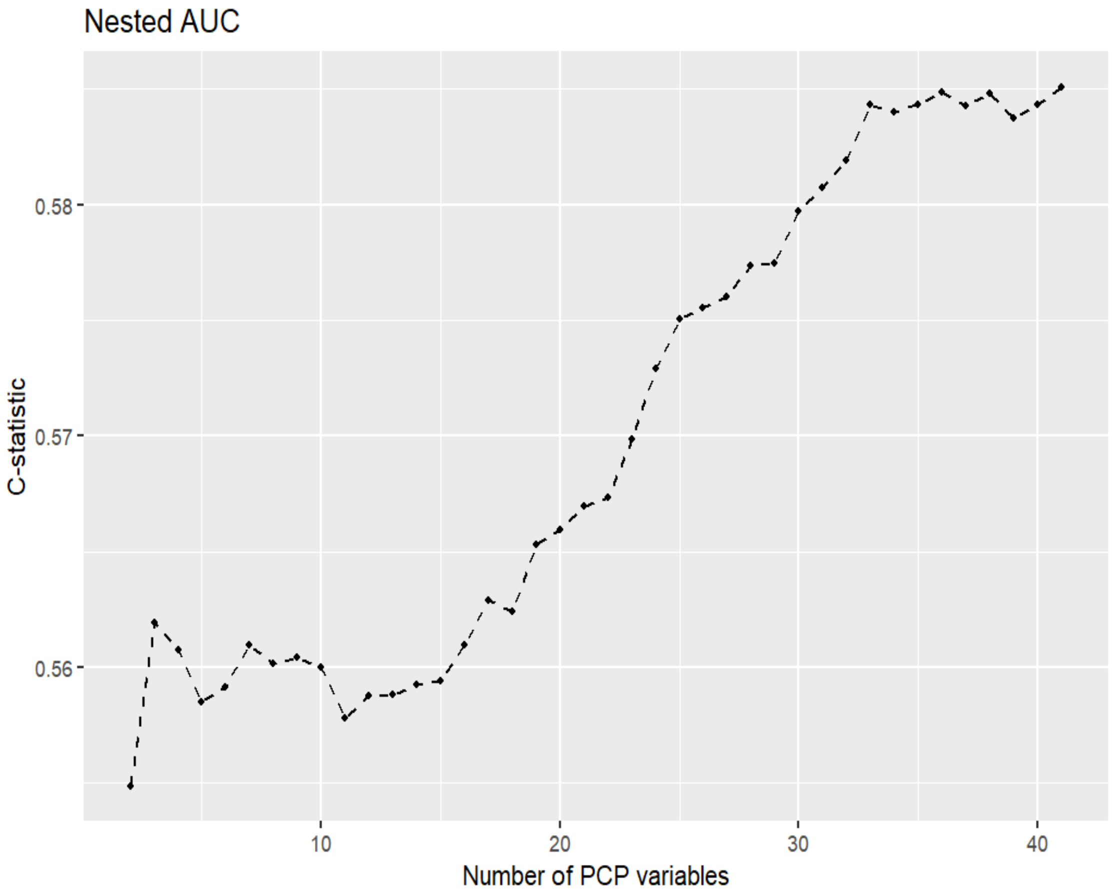

eTable 2. Summary of information for selecting number of latent classes of overall personal care product use [a]

| Number of Latent Classes | $G^2$ | AIC | BIC | CAIC | a-BIC |
|---|---|---|---|---|---|
| 2 | 1359975.66 | 1360505.66 | 1362715.67 | 1362980.67 | 1361873.50 |
| 3 | 1325381.50 | 1326177.50 | 1329496.68 | 1329894.68 | 1328231.84 |
| 4 | 1310596.15 | 1311658.15 | 1316086.50 | 1316617.50 | 1314398.99 |
| 5 | 1298709.27 | 1300037.27 | 1305574.80 | 1306238.80 | 1303464.61 |

G=Likelihood Ratio Chi-square; AIC=Akaike information criterion; BIC=Bayesian information criterion; CAIC=Consistent Akaike Information Criterion; a-BIC=sample size-adjusted Bayesian information criterion

[a] It is common for AIC to continue decreasing as the number of classes increases, which can lead to over-extraction. While BIC is generally more reliable, in cases of very large sample sizes, the penalty term can become too small relative to the log-likelihood, leading to over-extraction. Therefore, we considered not only the significant decrease in AIC and BIC values compared to the previous model, but also the substantive theory and interpretability to enhance the validity and interpretability of the model.

eTable 3. Latent class descriptions of overall personal care product use[a]

| Labels of latent classes | Class description |
|---|---|
| A. Infrequent users | Likely to have infrequent use of 33 personal care products (hair food, artificial nails (someone else), face cream, talcum powder applied vaginal area, artificial nails, douche, nail polish remover, body lotion, lip moisturizer, minoxidil, conditioner, lipstick, hand lotion, perfume, mascara, skin lightener, blush, nail polish, talcum powder applied other area, deodorant, talcum powder applied under arms area, pomade, eye liner, hair styling gel, hair spray, age spot lightener, baby oil, anti-aging product, self-tanning product, foundation, foot cream, cleansing cream, and bath gel) |
| B. Moderate users | Likely to have moderate use of hair food, talcum powder applied vaginal area, artificial nails, douche, body lotion, lip moisturizer, minoxidil, conditioner, lipstick, hand lotion, perfume, mascara, skin lightener, blush, nail polish, talcum powder applied other area, deodorant, talcum powder applied under arms area, pomade, eye liner, hair styling gel, hair spray, age spot lightener, baby oil, anti-aging product, self-tanning product, foundation, foot cream, cleansing cream, and bath gel, and likely to have frequent use of artificial nails (someone else), face cream, and nail polish remover |
| C. Frequent users | Likely to have frequent use of hair food, talcum powder applied vaginal area, artificial nails, douche, body lotion, lip moisturizer, minoxidil, conditioner, lipstick, hand lotion, perfume, mascara, skin lightener, blush, nail polish, talcum powder applied other area, deodorant, talcum powder applied under arms area, pomade, eye liner, hair styling gel, hair spray, age spot lightener, baby oil, anti-aging product, self-tanning product, foundation, foot cream, cleansing cream, and bath gel, and likely to have moderate use of artificial nails (someone else), face cream, and nail polish remover |

[a] Class labels and descriptions are based on likely item response probabilities for each product, but all responses for individual women may not fit these parameters; each class is described relative to the other classes in each product category (Taylor etal.2017).

eTable 4. Overall personal care product use and risk of hypertension stratified by race, menopausal status, and annual household income[a]

| | Infrequent users | Moderate users | Frequent users | P-trend | P for interaction[b] |
|---|---|---|---|---|---|
| | HR (95%CI) | HR (95%CI) | HR (95%CI) | | |
| Race and ethnicity | | | | | |
| White | 1 (Ref.) | 1.001 (0.95-1.06) | 1.08 (1.03-1.14) | 0.002 | 0.167 |
| Black | 1 (Ref.) | 1.13 (0.96-1.34) | 1.21 (0.97-1.50) | 0.082 | |
| Menopausal status | | | | | |
| Premenopausal | 1 (Ref.) | 0.997 (0.91-1.09) | 1.09 (1.001-1.19) | 0.034 | 0.937 |
| Postmenopausal | 1 (Ref.) | 1.03 (0.97-1.10) | 1.07 (1.01-1.13) | 0.033 | |
| Annual Household Income | | | | | |
| Less than $50,000 | 1 (Ref.) | 1.08 (0.98-1.19) | 1.12 (1.02-1.24) | 0.021 | 0.311 |
| $50,000 to $99,999 | 1 (Ref.) | 0.98 (0.91-1.07) | 1.06 (0.98-1.15) | 0.097 | |
| More than $100,000 | 1 (Ref.) | 0.99 (0.90-1.09) | 1.04 (0.95-1.14) | 0.343 | |
| Obesity Status | | | | | |
| Body mass index<30kg/m$^2$ | 1 (Ref.) | 1.01 (0.95-1.07) | 1.07 (1.01-1.14) | 0.011 | 0.723 |
| 30kg/m$^2$≤ Body mass index | 1 (Ref.) | 1.002 (0.92-1.10) | 1.06 (0.97-1.17) | 0.233 | |

HR=Hazard ratios; CI=95% confidence intervals; Ref.=reference group

[a] Models accounted for age as the timescale. In addition, models were adjusted for race/ethnicity (non-Hispanic Black, non-Hispanic White, Hispanic, other, or missing), body mass index (continuous, kg/m$^2$), education level (high school or less, some college, college and above, or missing), annual household income (<$50,000, $50,000-<$100,000, ≥$100,000), menopausal status at enrollment (premenopausal, postmenopausal, or missing), smoking pack-year (continuous, pack-year), alcohol consumption (never or past, current <1 drink, current ≥1 drinks, or missing), and physical activity (continuous, hours per week)

[b] Interactions were assessed through likelihood ratio tests by comparisons of Cox models with and without the cross-product of each factor and latent class.

eTable 5. Association between personal care products and risk of hypertension, additionally adjusting for family history of hypertension[a,b]

| | Infrequent users | Moderate users | Frequent users | Frequent users of talcum powder | P-trend |
|---|---|---|---|---|---|
| Product groups | HR (95% CI) | HR (95% CI) | HR (95% CI) | HR (95% CI) | |
| Beauty products | 1 (Ref.) | 0.999 (0.95-1.06) | 1.09 (1.04-1.15) | – | <0.001 |
| Everyday hair products | 1 (Ref.) | 0.98 (0.89-1.09) | 1.001 (0.90-1.12) | – | 0.603 |
| Hygiene products | 1 (Ref.) | 1.06 (1.00-1.13) | 1.12 (1.06-1.18) | 1.18 (1.10-1.27) | <0.001 |
| Skincare products | 1 (Ref.) | 1.02 (0.96-1.08) | 1.00 (0.95-1.05) | – | 0.939 |

HR=Hazard ratios; CI=95% confidence intervals; Ref.=reference group

[a] Numbers of total subjects and hypertension events are for women with complete data for each product class and covariates only

[b] Models accounted for age as the timescale. In addition, models were adjusted for race/ethnicity (non-Hispanic Black, non-Hispanic White, Hispanic, other, or missing), body mass index (continuous, kg/$m^2$), education level (high school or less, some college, college and above, or missing), annual household income (<\$50,000, \$50,000-<\$100,000, ≥\$100,000), menopausal status at enrollment (premenopausal, postmenopausal, or missing), smoking pack-year (continuous, pack-year), alcohol consumption (never or past, current <1 drink, current ≥1 drinks, or missing), physical activity (continuous, hours per week), and family history (biological parents) of hypertension

eTable 6. Association between frequency of individual beauty product use in the past 12 months before baseline and risk of hypertension[b]

| | Did not use | Less than once a month | 1-3 times per month | 1-5 times per week | More than 5 times per week | P-trend | FDR adjusted P-trend[c] |
|---|---|---|---|---|---|---|---|
| | HR (95%CI) | HR (95%CI) | HR (95%CI) | HR (95%CI) | HR (95%CI) | | |
| Blush or rouge | 1.00 (Ref.) | 1.00 (0.94-1.08) | 0.98 (0.91-1.06) | 1.02 (0.96-1.08) | 1.05 (0.998-1.11) | 0.047 | 0.077 |
| Eye liner | 1.00 (Ref.) | 0.99 (0.92-1.06) | 1.01 (0.93-1.09) | 0.97 (0.91-1.02) | 1.03 (0.97-1.08) | 0.667 | 0.667 |
| Eye shadow | 1.00 (Ref.) | 1.02 (0.96-1.08) | 1.02 (0.95-1.09) | 1.06 (0.998-1.12) | 1.11 (1.05-1.18) [a] | <0.001 | <0.001 |
| Foundation makeup | 1.00 (Ref.) | 1.05 (0.97-1.12) | 1.02 (0.95-1.11) | 1.08 (1.02-1.14)[a] | 1.12 (1.06-1.18) [a] | <0.001 | <0.001 |
| Lipstick | 1.00 (Ref.) | 0.98 (0.90-1.06) | 0.98(0.89-1.05) | 1.05 (0.98-1.12) | 1.08 (1.01-1.15) [a] | 0.001 | 0.002 |
| Eye mascara | 1.00 (Ref.) | 0.99 (0.92-1.06) | 1.05 (0.97-1.13) | 1.04 (0.97-1.10) | 1.07 (1.02-1.13) [a] | 0.004 | 0.007 |
| Perfume or cologne | 1.00 (Ref.) | 1.06 (0.99-1.13) | 1.07 (1.00-1.14) [a] | 1.12 (1.06-1.19)[a] | 1.18 (1.10-1.25) [a] | <0.001 | <0.001 |
| Makeup remover | 1.00 (Ref.) | 0.99 (0.93-1.06) | 1.03 (0.95-1.13) | 1.07 (1.003-1.15)[a] | 1.10 (1.03-1.16) [a] | 0.001 | 0.002 |
| Artificial nails/fill-ins someone else | 1.00 (Ref.) | 1.26 (1.02-1.56) [a] | 1.21 (0.89-1.65) | 0.63 (0.16-2.53) | - | 0.0890 | 0.129 |
| Artificial nails or fill-ins | 1.00 (Ref.) | 1.13 (1.04-1.23) [a] | 1.17 (1.09-1.25) [a] | 1.11 (0.70-1.77) | 0.74 (0.35-1.55) | <0.001 | <0.001 |
| Cuticle cream | 1.00 (Ref.) | 1.01 (0.96-1.05) | 1.06 (1.00-1.13) [a] | 1.04 (0.93-1.17) | 0.99 (0.76-1.28) | 0.112 | 0.146 |
| Nail polish | 1.00 (Ref.) | 1.04 (0.98-1.10) | 1.05 (0.99-1.11) | 1.01 (0.92-1.10) | 1.09 (0.91-1.32) | 0.326 | 0.385 |
| Nail polish remover | 1.00 (Ref.) | 1.03 (0.97-1.09) | 1.04 (0.98-1.10) | 0.96 (0.87-1.05) | 1.40 (0.94-2.08) | 0.666 | 0.667 |

HR=Hazard ratios; CI=95% confidence intervals; Ref.=reference group

[a] P values are less than 0.05.

[b] Models accounted for age as the timescale. In addition, models were adjusted for race/ethnicity (non-Hispanic Black, non-Hispanic White, Hispanic, other, or missing), body mass index (continuous, $kg/m^2$), education level (high school or less, some college, college and above, or missing), annual household income (<\$50,000, \$50,000-<\$100,000, ≥\$100,000), menopausal status at enrollment (premenopausal, postmenopausal, or missing), smoking pack-year (continuous, pack-year), alcohol consumption (never or past, current <1 drink, current ≥1 drinks, or missing), and physical activity (continuous, hours per week).

[c] The false discovery rate (FDR) adjustment was used to correct for multiple testing.

eTable 7. Association between frequency of individual everyday hair product use in the past 12 months before baseline and risk of hypertension[b]

| | Did not use | Less than once a month | 1-3 times per month | 1-5 times per week | More than 5 times per week | P-trend | FDR adjusted P-trend[c] |
|---|---|---|---|---|---|---|---|
| | HR (95%CI) | HR (95%CI) | HR (95%CI) | HR (95%CI) | HR (95%CI) | | |
| Hair conditioner/rinse | 1.00 (Ref.) | 0.94 (0.85-1.03) | 0.92 (0.84-1.00) [a] | 0.96 (0.90-1.02) | 0.95 (0.89-1.02) | 0.477 | 0.835 |
| Hair food | 1.00 (Ref.) | 1.07 (0.95-1.21) | 1.03 (0.88-1.22) | 0.94 (0.72-1.22) | 0.95 (0.53-1.37) | 0.966 | 0.966 |
| Hair spray | 1.00 (Ref.) | 1.03 (0.97-1.10) | 1.08 (1.01-1.16) [a] | 1.08 (1.02-1.14) [a] | 1.14 (1.07-1.20) [a] | <0.001 | <0.001 |
| Hair styling gel/mousse | 1.00 (Ref.) | 1.03 (0.96-1.10) | 0.97 (0.91-1.04) | 1.06 (1.003-1.12) [a] | 1.01 (0.95-1.08) | 0.258 | 0.602 |
| Minoxidil or Rogaine | 1.00 (Ref.) | 1.10 (0.79-1.54) | 0.91 (0.60-1.38) | 1.36 (0.99-1.87) | 0.86 (0.66-1.12) | 0.846 | 0.966 |
| Pomade or hair grease | 1.00 (Ref.) | 1.02 (0.92-1.13) | 1.02 (0.92-1.13) | 0.95 (0.86-1.06) | 0.79 (0.66-0.96) | 0.076 | 0.267 |
| Shampoo | 1.00 (Ref.) | 1.00 (0.51-1.97) | 0.97 (0.52-1.82) | 0.94 (0.51-1.75) | 0.96 (0.52-1.79) | 0.680 | 0.952 |

HR=Hazard ratios; CI=95% confidence intervals; Ref.=reference group

[a] P values are less than 0.05.

[b] Models accounted for age as the timescale. In addition, models were adjusted for race/ethnicity (non-Hispanic Black, non-Hispanic White, Hispanic, other, or missing), body mass index (continuous, $kg/m^2$), education level (high school or less, some college, college and above, or missing), annual household income (<\$50,000, \$50,000-<\$100,000, ≥\$100,000), menopausal status at enrollment (premenopausal, postmenopausal, or missing), smoking pack-year (continuous, pack-year), alcohol consumption (never or past, current <1 drink, current ≥1 drinks, or missing), and physical activity (continuous, hours per week).

[c] The false discovery rate (FDR) adjustment was used to correct for multiple testing.

eTable 8. Association between frequency of individual hygiene product use in the past 12 months before baseline and risk of hypertension[b]

| | Did not use | Less than once a month | 1-3 times per month | 1-5 times per week | More than 5 times per week | P-trend | FDR adjusted P-trend[c] |
|---|---|---|---|---|---|---|---|
| | HR (95%CI) | HR (95%CI) | HR (95%CI) | HR (95%CI) | HR (95%CI) | | |
| Bath or shower gel | 1.00 (Ref.) | 1.04 (0.98-1.10) | 1.07 (1.01-1.15) [a] | 1.05 (0.99-1.11) | 1.10 (1.04-1.16) [a] | 0.0015 | 0.002 |
| Deodorant and/or antiperspirant | 1.00 (Ref.) | 1.05 (0.87-1.27) | 0.88 (0.74-1.05) | 1.13 (0.995-1.28) | 1.27 (1.14-1.42) [a] | <0.001 | <0.001 |
| Douche | 1.00 (Ref.) | 1.11(1.04-1.19) [a] | 1.26 (1.11-1.42) [a] | 1.11 (0.78-1.59) | 1.29 (0.61-2.71) | <0.001 | <0.001 |
| Mouthwash/rinse | 1.00 (Ref.) | 1.03 (0.97-1.09) | 1.07(1.01-1.14) [a] | 1.07 (1.01-1.14) [a] | 1.11 (1.04-1.18) [a] | <0.001 | 0.001 |
| Shaving creams or gels | 1.00 (Ref.) | 1.07 (1.01-1.14) [a] | 1.11 (1.05-1.17) [a] | 1.05 (0.99-1.11) | 1.17 (1.02-1.34) [a] | <0.001 | 0.001 |
| Talcum powder under arms | 1.00 (Ref.) | 1.00 (0.92-1.07) | 1.16 (1.04-1.28) [a] | 1.15 (1.03-1.30) [a] | 1.05 (0.92-1.20) | 0.007 | 0.008 |
| Talcum powder on vaginal area | 1.00 (Ref.) | 1.07 (0.99-1.16) | 1.20 (1.08-1.33) [a] | 1.15 (1.02-1.30) [a] | 1.03 (0.90 -1.19) | 0.002 | 0.002 |
| Talcum powder to other areas | 1.00 (Ref.) | 0.99 (0.94-1.05) | 1.01 (0.94-1.08) | 1.03 (0.95-1.12) | 1.13 (1.03-1.24) [a] | 0.032 | 0.032 |

HR=Hazard ratios; CI=95% confidence intervals; Ref.=reference group

[a] P values are less than 0.05.

[b] Models accounted for age as the timescale. In addition, models were adjusted for race/ethnicity (non-Hispanic Black, non-Hispanic White, Hispanic, other, or missing), body mass index (continuous, $kg/m^2$), education level (high school or less, some college, college and above, or missing), annual household income (<\$50,000, \$50,000-<\$100,000, ≥\$100,000), menopausal status at enrollment (premenopausal, postmenopausal, or missing), smoking pack-year (continuous, pack-year), alcohol consumption (never or past, current <1 drink, current ≥1 drinks, or missing), and physical activity (continuous, hours per week).

[c] The false discovery rate (FDR) adjustment was used to correct for multiple testing.

eTable 9. Association between frequency of individual skincare product use in the past 12 months before baseline and risk of hypertension[b]

| | Did not use | Less than once a month | 1-3 times per month | 1-5 times per week | More than 5 times per week | P-trend | FDR adjusted P-trend[c] |
|---|---|---|---|---|---|---|---|
| | HR (95%CI) | HR (95%CI) | HR (95%CI) | HR (95%CI) | HR (95%CI) | | |
| Anti-aging/wrinkle products | 1.00 (Ref.) | 1.04 (0.97-1.11) | 1.08 (1.002-1.16) [a] | 1.03 (0.97-1.09) | 1.04 (0.99-1.09) | 0.143 | 0.316 |
| Age spot lightener | 1.00 (Ref.) | 1.12 (1.02-1.22) [a] | 0.94 (0.83-1.06) | 1.07 (0.95-1.19) | 1.06 (0.93-1.20) | 0.215 | 0.376 |
| Baby oil/mineral-based oils | 1.00 (Ref.) | 1.04 (0.97-1.10) | 1.11 (1.02-1.20) [a] | 1.00 (0.91-1.11) | 1.07 (0.96-1.19) | 0.061 | 0.284 |
| Blemish/acne products | 1.00 (Ref.) | 1.04 (0.97-1.10) | 1.07 (0.98-1.16) | 0.98 (0.88-1.08) | 1.00 (0.90-1.12) | 0.631 | 0.736 |
| Body lotions or creams | 1.00 (Ref.) | 1.03 (0.92-1.16) | 0.97 (0.87-1.07) | 0.98 (0.90-1.08) | 0.97 (0.89-1.06) | 0.263 | 0.409 |
| Cleansing cream | 1.00 (Ref.) | 1.02 (0.95-1.10) | 1.03 (0.95-1.11) | 0.98 (0.92-1.05) | 0.99 (0.94-1.04) | 0.487 | 0.682 |
| Face creams/moisturizers | 1.00 (Ref.) | 1.05 (0.94-1.18) | 0.97 (0.88-1.08) | 0.96 (0.89-1.04) | 0.96 (0.90-1.03) | 0.102 | 0.316 |
| Facial masks | 1.00 (Ref.) | 0.99 (0.95-1.04) | 1.01 (0.93-1.09) | 1.03 (0.87-1.23) | 0.87 (0.56-1.36) | 0.921 | 0.947 |
| Foot creams or moisturizers | 1.00 (Ref.) | 1.02 (0.96-1.08) | 1.05 (0.99-1.11) | 1.04 (0.98-1.11) | 1.09 (1.02-1.17) [a] | 0.007 | 0.094 |
| Hand lotions or creams | 1.00 (Ref.) | 1.12 (0.94-1.33) | 1.05 (0.90-1.24) | 1.06 (0.91-1.23) | 1.303 (0.88-1.19) | 0.158 | 0.316 |
| Lip moisturizers | 1.00 (Ref.) | 1.01 (0.94-1.09) | 0.97 (0.90-1.04) | 0.95 (0.89-1.02) | 0.95 (0.89-1.02) | 0.037 | 0.262 |
| Petroleum jelly | 1.00 (Ref.) | 0.96 (0.91-1.02) | 1.01 (0.93-1.09) | 0.93 (0.84-1.02) | 1.06 (0.97-1.16) | 0.947 | 0.947 |
| Skin lighteners | 1.00 (Ref.) | 1.14 (0.99-1.32) | 1.00 (0.83-1.20) | 0.97 (0.81-1.17) | 0.90 (0.73-1.11) | 0.606 | 0.736 |
| Self-tanning products | 1.00 (Ref.) | 1.04 (0.99-1.10) | 1.06 (0.98-1.14) | 1.05 (0.95-1.17) | 0.92 (0.69-1.23) | 0.115 | 0.316 |

HR=Hazard ratios; CI=95% confidence intervals; Ref.=reference group

[a] P values are less than 0.05.

[b] Models accounted for age as the timescale. In addition, models were adjusted for race/ethnicity (non-Hispanic Black, non-Hispanic White, Hispanic, other, or missing), body mass index (continuous, $kg/m^2$), education level (high school or less, some college, college and above, or missing), annual household income (<\$50,000, \$50,000-<\$100,000, ≥\$100,000), menopausal status at enrollment (premenopausal, postmenopausal, or missing), smoking pack-year (continuous, pack-year), alcohol consumption (never or past, current <1 drink, current ≥1 drinks, or missing), and physical activity (continuous, hours per week).

[c] The false discovery rate (FDR) adjustment was used to correct for multiple testing.
